# Reduction of Maternal Mortality: Intercultural Care for Pregnancy and Childbirth at the Hospital of Tartagal, Salta, Argentina

**DOI:** 10.1101/2025.05.07.25326956

**Authors:** Silvana Figar, Adrian Murillo, Monica Campos, Janet Meoniz, Emilse Palisa, Irma Quinteros, Carolina Rivarossa, Reina Sosa, Dario Taborga, Marcela Campero, Lara Agudin, Paula Zingoni, Gabriela Dorigato

## Abstract

**Objective:** To describe the factors influencing the reduction of maternal mortality during the 2022–2024 management period at Tartagal Hospital (HT).

**Methods:** Participatory Action Research (PAR) with a qualitative-quantitative analysis. Twelve meetings and interviews were conducted with the healthcare team, along with 10 validated surveys administered to obstetric inpatients (10/12/24), and an analysis of hospital databases.

**Results:** Since 2022, the maternal mortality rate at HT has been reduced to zero. The main contributing factor was improved accessibility through the implementation of intercultural care along the maternal care pathway and respectful childbirth for Indigenous communities, based on the Primary Health Care Program. A total of 59 Community Health Workers participated by monitoring communities, providing priority transportation for at-risk pregnant women, and conducting nutritional follow-ups in coordination with social programs. Five transcultural facilitators (Wichí and Guaraní) provided hospital support. Surveys showed a 90% compliance rate with Law 25.929 (Respectful Childbirth), with effective management of early mother-child contact. A need was identified to redesign wards, bathrooms, and lodging facilities for pregnant women from remote areas to strengthen this model.

**Discussion:** HT demonstrated that intercultural, comprehensive, and territory-based management of pregnancy and childbirth led to a reduction in maternal mortality beginning in 2022. Teamwork and intersectoral collaboration optimized available resources, consolidating a sustainable and effective model for maternal health. However, the greatest challenge lies in maintaining and increasing funding to ensure the continuity of this policy.

## Manuscript

The maternal mortality rate in the province of Salta, Argentina, for the year 2021 was 10.8 per 10,000 live births, placing it in the highest quartile among provincial rates in the country (min. 2.7 - max. 19). This figure dropped to 5.3 in 2022 and to 1.4 in 2023 (1). The Juan D. Perón Hospital (Tartagal Hospital), located in the city of Tartagal, is one of the main referral healthcare centers for the population in the northern region of the Salta province. This population exhibits some of the highest levels of socio-sanitary vulnerability in the country, with unmet basic needs reaching 46.11% in rural areas declared under socio-sanitary emergency (2).

Access to healthcare, as a citizen’s right to the resources that foster individual and collective development capacities, requires political and managerial integration across different levels of government. Additionally, it demands the guarantee of local managerial autonomy to address the specific social determinants that affect health care, disease management, and life processes within a particular territory (3)(4).

A significant proportion of the areas served by Tartagal Hospital are inhabited by rural Indigenous communities, posing additional challenges in terms of cultural and geographic accessibility to health care. According to preliminary data from the 2022 National Population Census, Salta’s total population was 1,440,672. A notable feature is its ethnic and cultural diversity: 6.5% of the population self-identify as descendants of Indigenous peoples, nearly triple the national average of 2.4%. Among this population, 57.4% live in urban or peri-urban areas and 42.6% in rural regions. At least sixteen Indigenous peoples currently self-identify across the province’s territory (2).

One key cultural tension involves differing worldviews on pregnancy care and childbirth. In response, Tartagal Hospital has strengthened its intercultural maternal care efforts. According to local statistics, since 2022 these efforts have helped reduce maternal mortality to zero among pregnant women within the hospital’s territorial responsibility.

This study, through a participatory action research (PAR) approach, analyzes the factors contributing to maternal mortality reduction during the 2022–2024 management period at Tartagal Hospital. It examines the characteristics, barriers, and facilitators in maternal care pathways and includes improvement proposals developed by local stakeholders. The goal is to systematize dimensions that enhance the intercultural management of pregnancy and childbirth (FIMEP).

Specifically, this study focuses on dimensions affecting access to prenatal and childbirth care at Tartagal Hospital and on support strategies for Indigenous populations. The study integrates multiple frameworks: the rights-based and intercultural approaches for symbolic accessibility (drawing on Law 25.929 on Respectful Childbirth) (5)(6); social determinants of health for geographic, economic, and administrative accessibility (7)(8); and complexity theory, systems thinking (9), and social change theory (10) to describe care transformation practices through active participation and critical reflection among Tartagal Hospital professionals and regional/national health ministry managers.

## Methodology

### Design

Participatory Action Research (PAR) (11) with qualitative-quantitative triangulation.

### Study setting

Juan D. Perón Hospital of Tartagal (Tartagal, Salta, Argentina), Level IV complexity, head hospital of Operational Area XII, with a 194-bed infrastructure. It serves the Chaco and Yungas regions across the departments of General San Martín, Rivadavia, Anta, and part of Orán. Indigenous groups include Wichí, Chorote, Chulupí, Iogys, Weenhayek, Tapiete, Qom (Toba), Guaraní, Tupí Guaraní, Ava Guaraní, and Chané. On average, rural communities travel 200 km to reach the hospital, which is the referral maternity center for hospitals in Santa Victoria Este, Salvador Massa, Alto la Sierra, Aguaray, and Mosconi. There are no health shelters (2).

### Data sources

#### Quantitative

- Annual report from the Primary Health Care Program, Operational Area XII Tartagal, Hospital J.D. Perón (2023)
- Health statistics from INDEC (1); Report for the International Bank for Reconstruction and Development by the University of Salta (2).
- Scale of practices and behaviors related to pregnancy and childbirth care, conducted on all hospitalized obstetric patients (n=10) on October 12, 2024. Instrument validated by the University of Huánuco – Peru, 2018 (12).

### Qualitative

- Records of 6 virtual meetings assessing political feasibility of technical assistance and interviews with central and local health managers.
- 12 interviews with healthcare staff regarding barriers and facilitators in maternal care, knowledge and attitudes toward Law 25.929, and identification of improvement cycles. The interview guide was based on Cruz Vega’s work (12) and the birth plan model of the Italian Hospital of Buenos Aires.

Qualitative materials were processed through content analysis (13) using Atlas-ti_2023 software. The Perodeau diagram (14) was used to model social determinants, and identified dimensions were reviewed with the PAR group.

### Ethical safeguards

The study stems from a management report conducted under the National Personal Data Protection Law 25.326. Oral, audio-recorded informed consent was obtained for voluntary participation and anonymized, confidential information. Use of audiovisual images was authorized through signed authorship consent.

## Results

### From the research process

- **Start of the PAR process: August 2024**. The Undersecretariat of Sectoral Relations and Coordination of the National Ministry of Health, in dialogue with the Undersecretariat of Social Medicine of the Ministry of Health of Salta, requested an epidemiological situational analysis, enabling political feasibility, the creation of a mixed research group, and planning of the fieldwork. This process was documented through an initial sociogram of 22 stakeholders (not detailed here for confidentiality) and an audiovisual product.
- **First PAR cycle: October 2024**. Technical assistance was carried out at the hospital to design the FIMEP. This included 12 interviews with healthcare staff, 1 focus group with all interviewees, and 10 surveys with hospitalized women in the obstetrics ward at Tartagal Hospital (HT).
- **Second PAR cycle: November 2024**. Dimensions of the determinants of accessibility to pregnancy and childbirth care were identified through iterative systems modeling, integrating qualitative and quantitative data. See Figure 1.

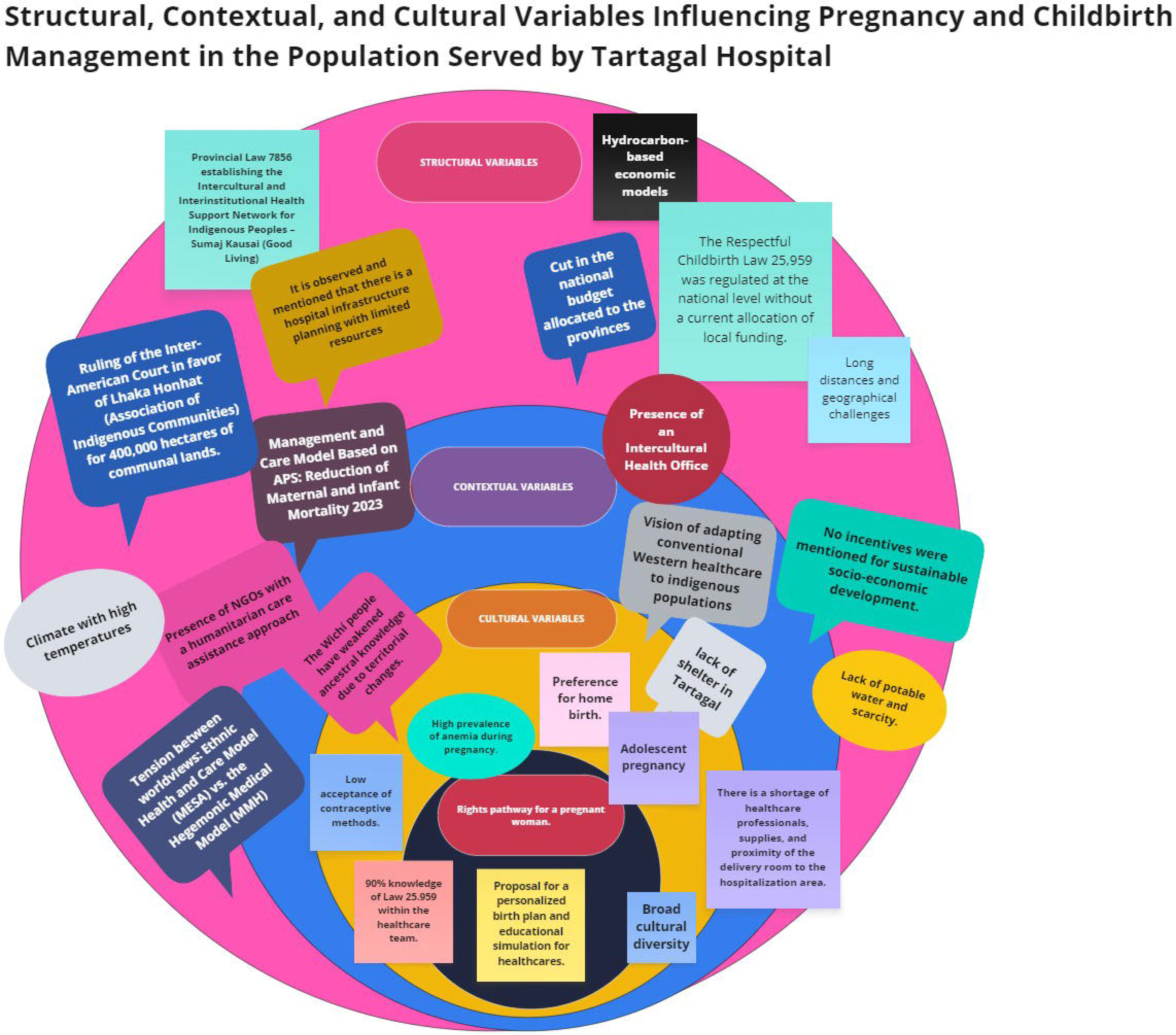
- **Third PAR cycle: December 2024**. Guidelines were developed for a Public-Private linkage mechanism, along with technical documentation for a communication plan of the Proposed Action Lines for Strengthening Intercultural Management of Pregnancy and Childbirth in Tartagal (FIMEP) See Table 1.

**Table 1:** Proposed Action Lines for Strengthening Intercultural Management of Pregnancy and Childbirth in Tartagal (FIMEP)

| **Action Line** | **Description** |
| --- | --- |
| **Intercultural Birth Plans** | Co-creation of birth plans with Indigenous communities, traditional midwives, and health professionals, respecting cultural practices. |
| **Digitalization and App** | Integration of birth plans into electronic health records and development of an app for access and monitoring. |
| **Transcultural Training** | Training healthcare personnel in intercultural approaches to improve maternal care. |
| **Integration into Pregnancy Monitoring** | Incorporation of sociocultural and biomedical information into the prenatal monitoring system. |
| **Public-Private Support Network** | Connecting pregnant women with resources and assistance through software that links needs with available support. |
| **Respectful Birth Simulation** | Video-recorded training sessions using simulation techniques to strengthen humanized care practices. |
| **Infrastructure Improvements** | Hospital adaptations for more humanized births: proximity to neonatology, family-friendly spaces, and better inpatient conditions. |
| **Global Citizen Participation** | Inclusion of the broader community in developing solutions to the social determinants of maternal care. |
| **Education and Implementation** | Systematization of the humanized birth model for replication in other populations and institutions. |
| **Health Information Systems (HIS)** | Development of bidirectional data systems for a comprehensive and contextual analysis of maternal health. |
| **Communication Strategy** | Use of social marketing, digital platforms, and media to raise awareness and promote participation and philanthropic donations. |

### Factors influencing the reduction of maternal mortality

Tartagal Hospital has a Primary Health Care (PHC) program with 59 territorial health agents (AS) and a family medicine residency program, both integrated into the Rural Program designed by Enrique Tanoni in 1978 (15). Since 2022, it also has an Office of Intercultural Affairs with five intercultural facilitators (Wichí and Guaraní).

In the interviews, **intersectoral collaboration** was highlighted as a key factor in reducing maternal mortality:

> “We have different institutions working on different issues, and we try not to operate in parallel. We hold regular meetings with the municipality and practice intersectorality. I think that’s what helped us avoid maternal mortality this year.”
>
> “…I think we didn’t have any maternal deaths because of how we work… we always ask how to reach high-risk cases. We always list them by name. We know the critical times of the year, like when diarrhea or heat leads to dehydration.”

### Key characteristics of the care and management model

#### Community follow-up and trust-building

Prenatal care begins directly in communities, especially in hard-to-reach rural areas.

> “During the community rounds, we identify pregnant women and children at risk of low weight. Then we organize a campaign. We already know that before the rains and dengue season, we must monitor underweight children. We call in low-weight cases and also take the opportunity to check the mothers.”

#### Early risk detection

By identifying risk factors in the pregnant woman’s home and environment—such as malnutrition, gender-based violence, or substance abuse—early interventions are designed.

> “In the last PHC round, 20% of families required prioritized follow-up due to risk factors.”

#### Multidisciplinary care

Health teams work in coordination with community psychology, gynecology and obstetrics, nutrition, the Intercultural Office, and social benefit services. This ensures more comprehensive, personalized care.

> “Having a multidisciplinary team opens several doors within the hospital to bring in mothers from remote areas, and provide them protected appointments and ultrasounds.”

#### Data recording and follow-up

The hospital has a data collection system to monitor pregnancies. Information is not only digitized but also displayed on large control panels in the meeting room, helping to visualize the pregnant women by territory.

#### Emotional and social support

Emotional support is provided to pregnant women and their families, particularly those in vulnerable situations.

> “They are accompanied by a community psychologist who runs empowerment activities in mother’s groups, addresses addiction, supports families who have lost children, and coordinates with local organizations using a network model.”

#### Intervention on social determinants of health

Health agents also report factors contributing to underweight children or unmonitored pregnancies. These include sexual abuse, gender or domestic violence, substance use, suicide attempts, and suicides. Community psychology services address these issues.

> “[Substance abuse] is a severe issue here due to the border situation; it used to not reach remote communities, but now it’s spreading…”

Problematic substance use cases are handled in coordination with the *Puente Norte Program*, part of the Mental Health and Addictions Secretariat of Salta. There was no mention of NGO support, but religious groups were noted as active collaborators.

#### Coordination with community leaders

Health agents work with tribal chiefs and ancestral midwives, strengthening ties with communities and ensuring culturally appropriate prenatal care.

#### Social motivation

Perhaps the most essential component of the management model is leadership that integrates the three key dimensions for transformative health management: symbolic, relational, and managerial (16).

> “We constantly ask ourselves: how do we keep going without losing the meaning of this work? How do we do good, dignified work for those who only have access to the public health system? Sometimes it’s overwhelming, but we have a team with a shared vision of public health. These conversations help us feel supported… Professional ethics guide us amid scarcity. But we’re tired. The needs are immense. It’s also a personal challenge—to stay focused on our goals and not get lost in the difficulties.”

Barriers and facilitators to prenatal and childbirth care are described across each dimension of accessibility.

### Geographical Accessibility

According to a report by the National University of Salta, the average distance from rural Indigenous communities to the hospital can reach up to 200 km (2).

The PHC management report states that transportation has been provided since 2020 through the Undersecretariat of Social Medicine, currently reinforced by the Indigenous Affairs Delegation—an effort shared by both departments.

> ““I had a patient who lived several kilometers away, across one of the rivers that’s difficult to cross. She was an adolescent in her third pregnancy—it would’ve been impossible for her to come on her own, so our entire team went to her.”“

Most interviewees referred to poor road conditions or the complete lack of roads, and the limited availability of public transportation in Indigenous-populated areas. There’s also a strong demand for increased funding for patient transportation. Currently, transportation is mainly used to move health teams.

> “What can’t be done at local health centers—ultrasounds, lab tests, specialist consults—sometimes doesn’t happen at the hospital either because of transport issues…”
>
> “They sometimes come in firewood trucks or vendor pickups. The roads are inaccessible.”
>
> “The institution does have transportation. It’s a coordinated effort, and we plan active outreach to ensure each pregnant woman receives quarterly check-ups, ultrasounds, and lab tests.”

The management report highlights that 50% of the motorcycles used by health agents (AS) for territorial visits are donated, allowing them to cover long distances and overcome geographic and climate barriers.

There was mention of a former shelter for distant families, which is seen as necessary—not only to mitigate transport issues but also to accommodate cultural norms of group movement.

### Administrative Accessibility

This refers to the availability of healthcare services, infrastructure, and human resources, including the number and quality of health centers, professionals, medications, and equipment.

- There are 13 health centers and 2 small satellite clinics (Tonono and Pacara, for more remote areas).
- In Km3 and Km6—more densely populated zones—the family medicine residency program supports care, with 2 residents, 2 staff doctors, and one instructor.
- In the fourth PHC round of 2023, there were 54 health agents, 7 supervisors, 1 administrator, and 1 program head. They provided access to 7,734 families across 58 out of 118 health sectors, with 1,500 families prioritized due to risk factors.

The local team emphasized the importance of increasing the number of health agents, with better pay, to improve territorial coverage.

Interviewees also requested hospital infrastructure improvements: Closer proximity between the delivery room, neonatology, and inpatient ward,”Home-like” spaces, Family shelters and improvements to the inpatient maternity ward.

> “We need maternity ward maintenance, replacement of broken furniture, bathroom repairs so patients can bathe. The mattresses get soiled with lochia and deteriorate, and there are no replacements… Since this hospital opened, it’s never been upgraded. The elevators are slow freight elevators, constantly broken…”

Another key issue is **finding specialists** for complicated births.

> “In Santa Victoria and Alto de la Sierra, there are general practitioners for monitoring, but births have to be referred to here.”

A critical enabling factor for Tartagal’s team is a sense of **“belonging”** to the place:

> “We need more doctors. Many retired and weren’t replaced. Doctors from the capital resist coming here…”

The interviews also revealed generational gaps in how the medical profession is valued, with a huge discrepancy between the effort required and the economic outlook of essential vs. popular specialties.

> “Fewer young people choose frontline specialties (e.g., internal medicine, gynecology), and even fewer are willing to work in remote areas.”
>
> “It’s not enough to just have a vocation or a dream.”
>
> “The biggest injustice is the wage gap between specialties… it’s a structural problem across the country… someone has to change that.”

The interviews pose broader questions for the national healthcare system: How can such a model be sustained if the next generation won’t continue as self-sacrificing “hero doctors”?

Regarding supplies, the list of shortages includes everything from ultrasound machines to blood bags:

> “That forces us to hospitalize patients who could have been treated with a transfusion—wasting inpatient resources.”

### Symbolic Accessibility

All interviews praised the recent creation of the Intercultural Health Office, which employs five transcultural facilitators (FTC)—Wichí and Guaraní—one of whom is a traditional midwife working in obstetrics.

In surveys regarding Law 25.929 (Respectful Childbirth Law), 90% of inpatients positively rated the care they received. The obstetric team, supported by the neonatal service, was engaged in a quality improvement cycle to strengthen early and continuous newborn contact.

There’s a clear conceptual gap between biomedical and Indigenous understandings of health:

> “Indigenous people don’t view health as we do. They don’t recognize the importance of prenatal or child checkups. We have illiterate patients, pregnant girls under 16, and lifelong undignified living conditions—no access to potable water.”

Facilitating birth plans under the Respectful Birth Law requires

### intercultural mediation

> “We sometimes mistake their silence for apathy, but it’s not that—they just don’t perceive health like we do. A healthy baby takes great effort…”

As with other hospitals, there’s a need for self-awareness around institutional culture, often described as the Hegemonic Medical Model (HMM) (17):

> “Everyone knows the respectful birth law. What they don’t get is that **they’re not the protagonists**—the mother is. That’s hard to internalize and defend…”
>
> “There’s a clash between **hegemonic, often macho medical culture** and community perspectives. Many fear coming due to past mistreatment. I’ve seen this firsthand. That’s why the presence of **intercultural facilitators helps tremendously**.”

**Empowerment of Indigenous women** is visible: female caciques helped establish the local health center.

A gender perspective is present among most female health workers, evident in how they explain, educate, and act:

> “It’s cultural for women to become mothers very young. They often reject contraceptives. We work on this at school too. It took a long time to get them to accept transdermal patches.”

Despite initial resistance, trust built over time has led to acceptance of contraception, which likely contributed to reduced teen pregnancies.

> “They now accept contraception—before, they didn’t even want to talk about it. They have access to condoms, but culture prevents their use. Still, it’s taught in schools.”

During technical assistance, a city-based birth plan was reviewed and adapted for local, intercultural use.

> “Women here struggle to express themselves. They might seem to understand during prenatal visits, but it’s not a conversation—they listen silently and come back with questions next time.”
>
> “They want to make their point clear—at the next appointment. We need a very tailored approach because each ethnic group is different. It’s usually the husband who comes and asks for birth control.”

#### “They want home births”

In Indigenous cultures, childbirth far from the community is not customary. Through empathetic listening during prenatal care, women can often be convinced to come to the hospital, but each case requires enormous effort.

> “We can inform them about the benefits of institutional births and prenatal care—but it’s hard. I might tell you to take iron supplements, but I can’t give it to you, or you have to come get it.”
>
> “It’s not just about putting a woman in an ambulance—maybe she has other children she can’t leave…”

### Economic Accessibility

Across all interviews, Argentina’s current economic crisis emerged as an overwhelming determinant, with a shared perception that the system is operating in “alert mode” and on the brink while managing a highly complex issue.

> “The thing is, a child in adverse conditions, now with 40°C temperatures, is at greater risk. We have to stay alert and focused. We’ve normalized the routines—how often to weigh them, when to refer—but in parallel we also have to build support networks in the healthcare system. It’s very hard to sustain all of this.”
>
> “We feel like we’re right on the edge. It’s a constant need to resolve things. We work a lot as a team to solve each particular case. Our answer to the people is always one that doesn’t involve saying there’s no solution here.”

## Discussion

The comprehensive management model built upon the altruistic commitment of the healthcare team has enabled a significant reduction in maternal mortality at the Tartagal Hospital. Teamwork capacity and sensitivity to patient needs have been key success factors. Moreover, intersectoral coordination and networked continuity of individualized care are also central components of this model.

The findings of this research align with the strategy recently published by the Pan American Health Organization (PAHO), which proposes that in order to reduce maternal mortality, care models must be centered on women, families, and communities (18).

Allocating resources for the implementation of sexual and reproductive health policies is an urgent requirement. However, the effectiveness of these policies will depend on innovative approaches to tackling this complex issue. The model promoted by PAHO places women not only as recipients, but as social innovators and decision-makers. Society could benefit from the differentiated capacities offered by gender and care perspectives, which help to evolve traditional relational models historically rooted in dominance and control (16).

However, in Argentina, there has recently been a withdrawal of support for these policies by the national government, shifting the entire burden to the provinces. This has not contributed to reducing inequalities in access to essential services—especially in the poorest provinces and those with larger Indigenous populations.

Our results are consistent with studies conducted in rural populations of northern Argentina, both in describing barriers to healthcare access and in highlighting the value of the integrated primary health care (PHC) model as a way to address them (19).

The literature also notes similar barriers in achieving respectful childbirth, particularly the cultural challenges posed by the hegemonic medical model (17). In Colombia, a study found that women have internalized mistreatment, making it essential for training on the Respectful Childbirth Law to adopt multiple strategies. These include promoting written birth plans as legal documents of the pregnant woman’s will, alongside training and simulations that help change practices and ultimately shift cultural norms to eliminate obstetric violence (20).

This study shows that the Tartagal Hospital health team has the skills to produce video simulations of the respectful childbirth process. With appropriate funding articulation (as outlined in Table 1), professional creativity could make it possible to disseminate and educate through an integrated strategy, grounded in a PHC-based management model that has already proven successful in reducing maternal mortality.

## Data Availability

All data produced in the present work are contained in the manuscript

## Acknowledgments

To the healthcare team and the hospitalized pregnant women who kindly contributed to this research.

